# Plasma pTau-217 in preclinical Alzheimer’s disease

**DOI:** 10.1101/2022.06.09.22276206

**Authors:** Erin M. Jonaitis, Shorena Janelidze, Karly A. Cody, Rebecca Langhough Koscik, Lianlian Du, Nathaniel A. Chin, Niklas Mattsson-Carlgren, Kirk J. Hogan, Bradley T. Christian, Tobey J. Betthauser, Oskar Hansson, Sterling C. Johnson

## Abstract

**Background and Objectives:** An accurate blood test for Alzheimer’s disease (AD) that is sensitive to preclinical proteinopathy and cognitive decline has clear implications for early detection and secondary prevention of AD. We assessed the performance of plasma pTau against brain PET markers of amyloid ([^11^C]-PiB) and tau ([^18^F]MK-6240), and its utility for predicting longitudinal cognition.

**Methods:** Samples were analyzed from a subset of participants with up to 8 years follow-up in the Wisconsin Registry for Alzheimer’s Prevention (WRAP; 2001-present; plasma 2011-present), a longitudinal cohort study of adults from midlife, enriched for parental history of AD. Participants were a convenience sample who volunteered for at least one PiB scan, had usable banked plasma, and were cognitively unimpaired at first plasma collection. Study personnel who interacted with participants or samples were blind to amyloid status. We used mixed effects models and receiver-operator characteristic curves to assess concordance between plasma pTau_217_ and PET biomarkers of AD, and mixed effects models to understand the ability of plasma pTau_217_ to predict longitudinal performance on WRAP’s preclinical Alzheimer’s cognitive composite (PACC-3).

**Results:** The primary analysis included 165 people (108 women; mean age=62.9 ± 6.06; 160 still enrolled; 2 deceased; 3 discontinued). Plasma pTau_217_ was strongly related to PET-based estimates of concurrent brain amyloid (β̂_*DVR*_ = 0.83 (0.75, 0.90), p<.001). Concordance was high between plasma pTau_217_ and both amyloid PET (AUC=0.91, specificity=0.80, sensitivity=0.85, PPV=0.58, NPV=0.94, *LR*^−^=5.48) and tau PET (AUC=0.95, specificity=1, sensitivity=0.85, PPV=1, NPV=0.98, *LR*^−^=6.47). Higher baseline pTau_217_ levels were associated with worse cognitive trajectories (β̂_*pTau=age*_ = -0.07 (-0.09, -0.06), p<.001).

**Conclusions and Relevance:** In a convenience sample of unimpaired adults, plasma pTau_217_ levels correlate well with concurrent brain AD pathophysiology and with prospective cognitive performance. These data indicate that this marker can detect AD before clinical signs and thus may disambiguate presymptomatic AD from normal cognitive aging.

**Classification of Evidence:** This study meets Class III evidential criteria for diagnostic accuracy of plasma pTau_217_.

## Introduction

Blood-based biomarkers for Alzheimer’s disease (AD) that detect beta-amyloid (A*β*) and phosphorylated tau (pTau) proteinopathy are rapidly developing.^1, 2^ The utility and convenience of an accurate blood test has clear implications for accelerating and improving clinical research and practice.^1–4^ Several candidate markers exist including mass-spectrometry^5–7^ and immunoassay^8^ measured A*β*_42_ and A*β*_40_ and their ratio, and phosphorylated tau at threonine 217 (pTau_217_),^9^ 181 (pTau_181_),^10^ and other phosphorylated sites,^11^ as well as non-specific markers of neurodegeneration and astrogliosis, including neurofilament light (NfL)^12, 13^ and glial fibrillary acidic protein (GFAP).^14–16^

Recently, interest has turned to pTau_217_, as cerebrospinal fluid levels increase early in autosomal dominant AD^17^ and better discriminate AD from non-AD subgroups of cognitively impaired adults, compared to pTau_181_.^18^ In plasma, pTau_217_ accurately differentiates persons with neuropathologically defined AD from other dementia.^9, 19^ Further, in vivo plasma pTau_217_ levels correlate with ex vivo protein levels and spatial burden in post-mortem brain tissue.^19–21^ Next, plasma pTau_217_ levels discriminate diagnostic groups informed by amyloid PET. pTau_217_ levels are elevated among impaired (AD or mild cognitive impairment (MCI)) A*β*^+^ participants compared to cognitively unimpaired (CU), A*β*^−^ participants,^11, 19^ and plasma pTau and tau PET signal show moderate to high agreement.^9, 11, 22^ Serial plasma pTau_217_ levels also differentiate AD from non-AD MCI, remaining stable and non-elevated in A*β*^−^ patients, and increasing over time in A*β*^+^ patients.^23^

The utility of plasma pTau_217_ to identify amyloid and tau proteinopathy in a preclinical cohort is less well studied. Among older adults in the Swedish BioFINDER study (mean age=72), pTau levels increased over 6 years in A*β*^+^ CU, but not A*β*^−^,^23^ similar to findings in MCI. In this same cohort, baseline pTau_217_ levels affected cognitive change.^3^ In the Australian Imaging, Biomarker & Lifestyle study (AIBL), among CU adults (mean age=75), a twofold increase in levels of the pTau + marker in A*β*^+^ compared to A*β*^−^ was recently reported,^11^ although the correlation between this biomarker and A*β* centiloids was relatively weaker in CU than in AD and MCI, perhaps due to restriction of range (ρ = 0.64 vs. 0.45). In the Mayo Clinic Study of Aging, among CU adults (mean age=79), a smaller fold increase of 0.49 was reported in A*β*^+^ compared to A*β*^−^.^24^ Measurement precision may depend in part on the instrument platform and assay.^18, 25, 26^

Here we report a study from the Wisconsin Registry for Alzheimer’s Prevention (WRAP)^27^ in which we examine plasma pTau_217_ trajectories in CU adults using Lilly’s immunoassay for the Meso Scale Discovery platform (Lilly-MSD) [palmqvist_discriminative_2020]. For this study, participants had a mean age of 63 at first plasma collection. We examined A) whether changes in plasma pTau_217_ levels over time track progression of AD proteinopathy ascertained from amyloid and tau positron emission tomography (PET) with [^11^C]-PiB for amyloid and [^18^F]MK-6240 for tau; B) whether plasma pTau_217_ levels accurately differentiate people with varying degreess of amyloid and tau burden; and C) whether plasma pTau_217_ levels are associated with longitudinal cognition.

## Methods

### Ethics

The research protocol was approved by the University of Wisconsin-Madison Health Sciences IRB (IRB00000366), and all participants provided written informed consent.

### Participants

Plasma samples were analyzed from WRAP participants with ≥ 1 amyloid PET scan using Pittsburgh Compound B (“PiB”; see Imaging methods). Participants were included in the PiB+ sample if they had ≥ 1 global PiB distribution volume ratio (DVR) > 1.19 (centiloid equivalent = 21.6). The PiB- sample included all participants who had ≥ 2 PiB scans with all global DVR ≤ 1.1. We also examined samples from participants whose global PiB DVR trajectories indicated possible conversion from PiB- to PiB+ by virtue of initially low but recently subthreshold DVR values (1.16 < DVR ≤ 1.19). Primary analyses included only participants who were cognitively unimpaired (CU) at their first plasma collection, and excluded one participant whose levels of pTau_217_ were highly influential in models.

Secondary analyses were conducted including these excluded participants (sensitivity set 1), and excluding participants with measured pTau_217_ below the lower limit of detection (sensitivity set 2; see Plasma methods and Supplement).

### Plasma methods

30 mL of blood were drawn from each participant into 3 × 10 mL lavender-top EDTA tubes (BD 366643; Franklin Lakes, New Jersey, United States). Samples were mixed gently by inverting 10-12 times and were centrifuged 15 minutes at 2000g at room temperature within 1 hour of collection. Plasma samples were aliquoted into 2 mL cryovials (Wheaton Cryoelite W985863; Millville, New Jersey, United States). Aliquoted plasma was frozen at -80 °C within ninety minutes and stored for up to 10 years.

Plasma pTau_217_ concentration was measured at the Clinical Memory Research Unit, Lund University (Sweden) using immunoassay on a Meso Scale Discovery (MSD) platform developed by Lilly Research Laboratories.^9^ Samples were assayed in duplicates according to published protocols^28^ with biotinylated-IBA493 used as a capture antibody and SULFO-TAG-4G10-E2 as the detector. The assay was calibrated with a synthetic pTau_217_ peptide. The mean intra-assay coefficient of variation (CV) was 7.11%. The inter-assay CV for 3 quality control samples included in every run was 10.3%. Plasma pTau_217_ concentration was below the detection limit of the assay (0.11-0.17 pg/mL) for 6 cases. For each model, sensitivity analysis 2 excluded these observations. Samples were arranged on plates according to a randomization scheme devised by author EMJ, who had no contact with samples. All samples were analyzed by staff blind to clinical and imaging data.

### Imaging methods

Participants underwent T1-weighted magnetic resonance imaging as well as amyloid ([^11^C]-PiB) and tau ([^18^F]-MK-6240) PET imaging at the University of Wisconsin-Madison. Detailed methods for radioligand synthesis and PET and MRI acquisition, processing and quantification, and analysis were implemented as reported previously.^29, 30^

Amyloid burden was assessed as a global cortical average [^11^C]-PiB DVR, and two DVR thresholds were applied for determining PiB positivity (PiB+): one at DVR > 1.19, based on previously published work,^31^ and another, lower threshold of DVR > 1.16 (corresponding to a centiloid of 17.7), previously shown to predict subsequent accumulation of amyloid.^32^ Estimated age of amyloid onset (EAOA) was obtained from observed global Pib DVR using a combination of group-based trajectory modeling and Bayes’ theorem^33^ using either the most recent PET scan (for those who were PiB-) or the scan closest to the PiB+ threshold. Amyloid duration was then estimated as age at plasma sample minus EAOA and the corresponding estimated PiB DVR (GBTM-DVR) was calculated via linear transformation as described in Betthauser et al.^34^ Centiloids were estimated from these DVRs according to the following equation: *CL* = 148.33 × *DVR* − 154.96.

[^18^F]-MK-6240 standardized uptake volume ratio (SUVR) (70-90 minutes; cerebellum gray reference region excluding the superior medial vermis) tau burden was assessed visually by an expert reader (SCJ) using SUVR images overlaid on the coregistered MRI and scaled from 0-2.5. Images were classified as tau negative or tau positive for the medial temporal lobe (MTL; entorhinal cortex, amygdala or hippocampus) and for the neocortex (Neo; 1 or more cortical regions). The visual rating defined four classes: MK-, MK+ in MTL only, MK+ in neocortex only, and MK+ in MTL and neocortex.

### Neuropsychological assessment and cognitive status

Participants in WRAP completed a comprehensive cognitive battery at each visit, including tests of memory, executive function, language ability, and other aspects of cognitive performance, alongside self- and informant-based measures of everyday functioning.^27^ Based on these measures, participant cognitive status at each visit was determined via consensus conference.^35^ Among those without clinically significant cognitive impairment (i.e., dementia or MCI), some were assigned a research diagnosis of “Cognitively Unimpaired-Declining” denoting performance within the range of normal, but suggestive of decline from baseline.^36^

Our measure of global cognition was a three-test version of the Preclinical Alzheimer’s Cognitive Composite^37^ including the Rey Auditory-Verbal Learning Test, sum of Trials 1-5; the Wechsler Memory Scale Logical Memory II, total score; and the Wechsler Adult Intelligence Scale-Revised Digit Symbol Substitution, total score. Tests were combined by rescaling and computing an unweighted average, scaled such that first observations in cognitively unimpaired indiduals were distributed ∼ *N*(0,1). The Wide-Range Achievement Test-Reading Subtest (standard score) was used as a measure of literacy.

### Statistical analysis

Statistical analyses were performed in R 4.0.5.^38^ Longitudinal pTau_217_ trajectories were modeled using mixed effects models^39^ with participant-level random intercepts, which are robust to missingness when data are missing at random. Two such models were fit. First, to evaluate how well pTau_217_ measurements reflect brain amyloid, we estimated the fixed effect of GBTM-DVR at each timepoint. Second, to compare age trends in people known to be accumulating amyloid versus people who are not,^23^ we estimated the fixed effects of age, amyloid status (PiB+ vs PiB-), and their interaction. Effect sizes were estimated by *Ω*^2^,^40^ where 0.01 ≤ *Ω*^2^ < 0.06 was considered a small effect size, 0.06 ≤ *Ω*^2^ < 0.14 medium, and *Ω*^2^ ≥ 0.14 large.^41^ Test-retest reliability was assessed using the intraclass correlation.

To establish potential thresholds with maximal correspondence between pTau_217_ and binary brain amyloid (global PiB) and tau (MTL+neocortical MK-6240) positivity, receiver-operator characteristic (ROC) curves^42^ were constructed on a sub-sample comprising 1 plasma observation per participant, acquired within 2 years of a PiB or MK-6240 scan, respectively. Thresholds were selected to maximize Youden’s index.^43^ Positive and negative predictive values for PiB- and MK-positivity assumed population prevalence of 25% and 10%, respectively. In a secondary analysis, we used a robust norms approach to identify an alternate threshold for pTau217 by first winnowing the sample to solidly PiB- individuals (DVR < 1.1 at all scans); computing the 2.5th and 97.5th percentiles; selecting all observations within this range; and recomputing the 97.5th percentile to obtain the robust-norms threshold. To validate these thresholds, we classified GBTM-DVR at each plasma observation into PiB- (GBTM-DVR ≤ 1.19) and PiB+ (GBTM-DVR > 1.19), and compared this against positivity on pTau_217_ according to each threshold.

To evaluate the relationship between baseline pTau_217_ and cognitive trajectories, we fit a mixed effects model of longitudinal PACC-3, with linear and quadratic age terms and their interaction with baseline pTau_217_ modeled as continuous fixed effects, and a participant-level random intercept. Age and pTau_217_ terms were mean-centered. Sex, education, baseline literacy, and number of prior exposures to the battery were included as covariates. For comparison, a covariates-only model was also fit.

To assess whether within-person change in pTau_217_ predicts within-person change in cognition, and explore the phasing of this relationship, an exploratory, repeated measures correlation analysis was performed.^44, 45^ This analysis used a subset of data in which pTau_217_ was paired variously with concurrent PACC-3 scores and with PACC-3 lagged by one or two study visits. Only participants with at least four timepoints were included (N=46; N_obs_ = 93), to satisfy the constraints that each participant should contribute at least two pTau_217_ observations, to assess within-person change, and that each such observation should allow for pairings with cognition under three lag conditions (e.g., pTau_217_ at Visits 1 and 2 with PACC-3 at (A) Visits 1 & 2, (B) Visits 2 & 3, and (C) Visits 3 & 4, in successive models lag=0, lag=1, lag=2). This exploratory analysis was repeated twice, first using PACC-3 and pTau_217_ values from which age had been partialed out, and then substituting GBTM-DVR for pTau_217_.

### Data availability

Coded data and R scripts underlying all analyses may be shared at the request of any qualified investigator for purposes of replication.

## Results

### Participants

173 participants had qualifying PiB scans and at least one plasma sample stored in EDTA. At their last PiB scan, 74 had PiB DVR > 1.19, and 99 had PiB DVR ≤ 1.19. Included in the second set were 84 having ≥ 2 PiB scans with all DVR ≤ 1.11, and 15 with values suggestive of possible conversion. From this set, 8 participants were removed from primary analyses due to cognitive impairment at first plasma (N=7), missing diagnosis at first plasma (N=1), or measured pTau_171_ levels found to be highly influential (N=1).

Participant characteristics for the primary analysis sample are shown in Table 1. For each applicable aim, excluded participants were included in a sensitivity analysis (sensitivity set 1; see eTable 1 and eFigures 1 and 3). 6 participants had pTau_171_ levels below the lower limit of detection; these were excluded for a second sensitivity analysis (sensitivity set 2; see eTable 1 and eFigures 2 and 4).

**Figure 1.**
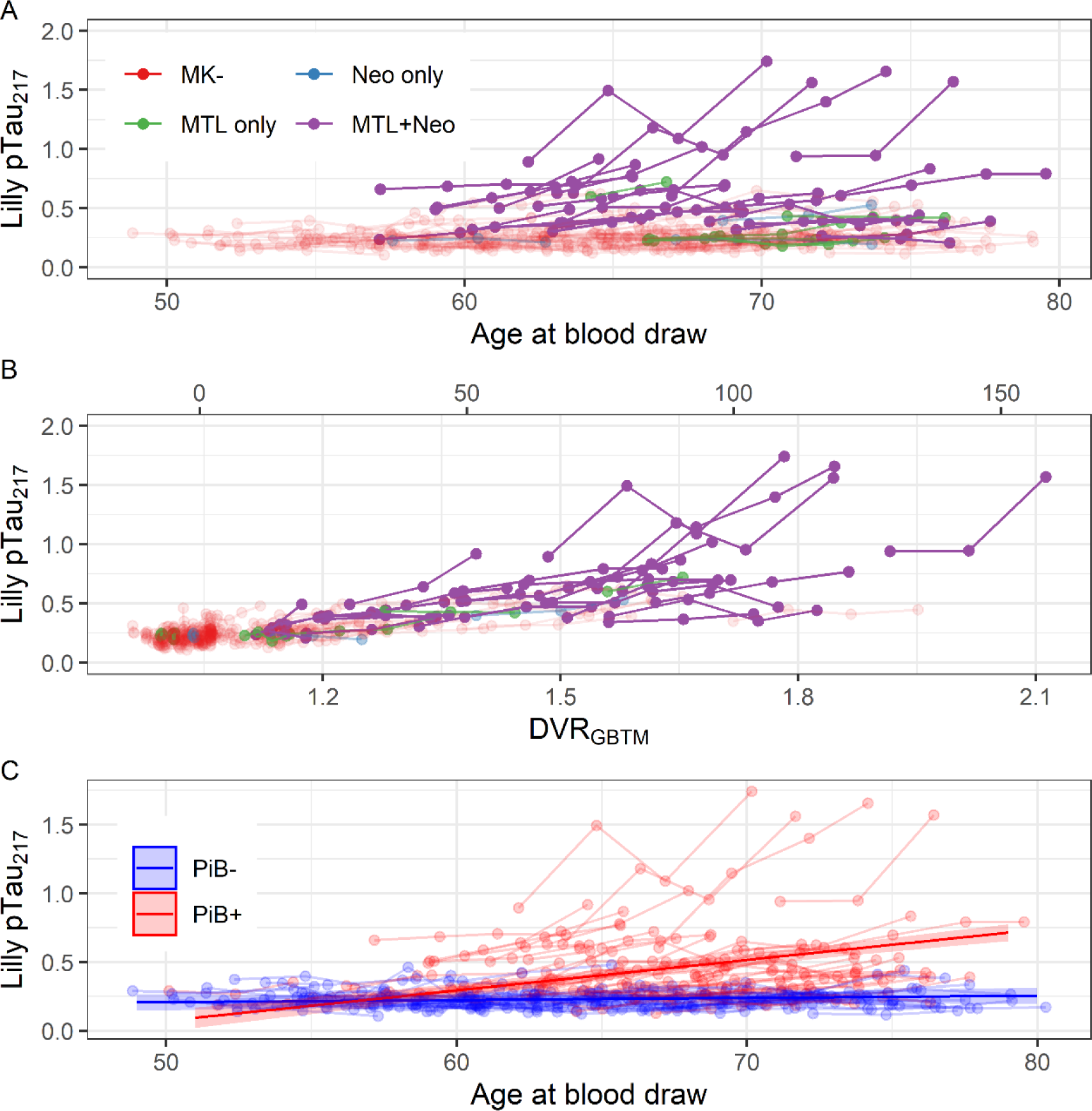
Longitudinal plasma pTau217. Observations from a single participant are shown with connected edges. (A) Plasma pTau217 as a function of age at blood draw. Color indicates the extent of tau burden as indicated on tau PET. (B) Plasma pTau217 as a function of estimated PiB DVR at the time of plasma acquisition. Color indicates the extent of tau burden as indicated on tau PET. (C) Plasma pTau217 as a function of age at blood draw. Color indicates amyloid PET positivity. Lines with shaded confidence bands represent slope estimates from a linear mixed effects model of pTau217 as a function of the interaction of age and amyloid positivity.

**Figure 2.**
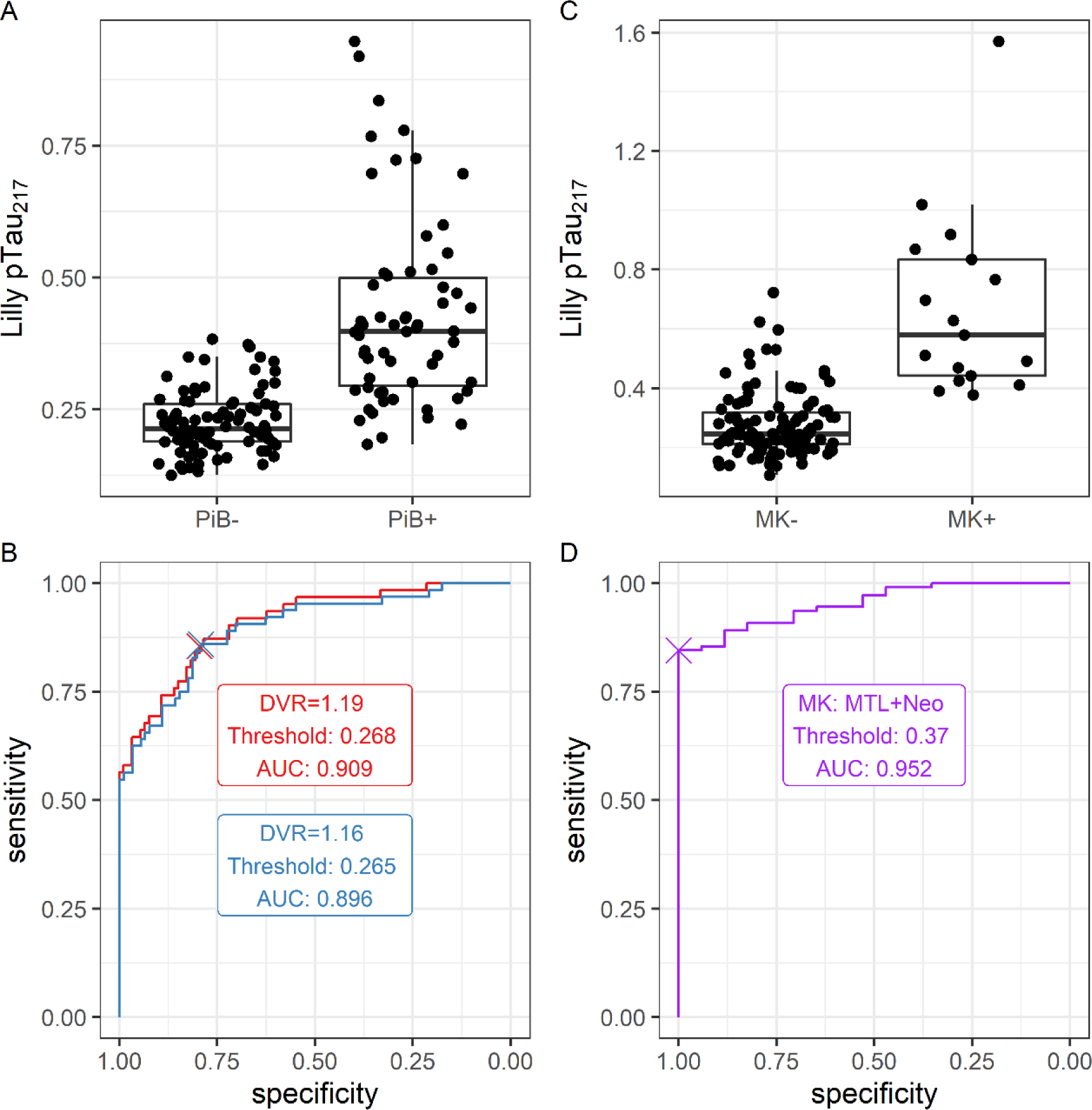
Relationship between pTau217 and PET AD biomarkers. (A) Distribution of pTau217 among PiB- and PiB+ participants. (B) Receiver-operator characteristic (ROC) curve relating pTau217 to binary PiB status. Two positivity thresholds were considered for PiB: global DVR > 1.19 (red) and global DVR > 1.16 (blue). (C) Distribution of pTau217 among MK- and MK+ participants. (D) ROC curve relating pTau217 to binary MK status. Scans were marked as MK+ if tracer binding was evident in both medial temporal lobe and neocortex, and MK- otherwise.

**Figure 3.**
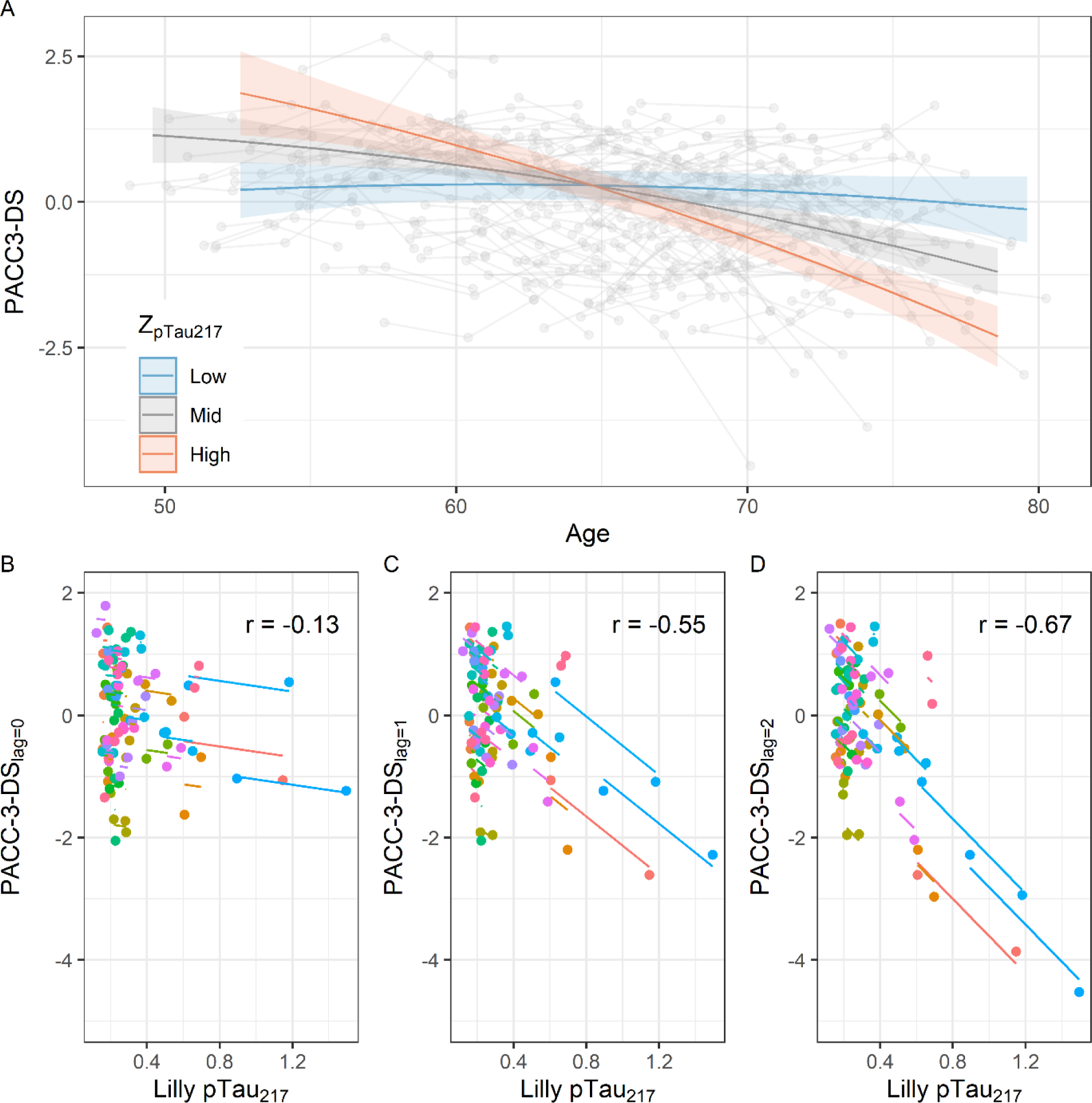
Relationships between pTau217 and longitudinal cognition. (A) Global cognition (PACC-3) as a function of age and baseline pTau217 level. Low, medium, and high pTau217 values reflect the 10th, 50th, and 90th sample percentiles. (B-D) Repeated measures correlation between global cognition (PACC-3) and pTau217 with lags of 0 (B), 1 (C), and 2 (D) visits between biomarker and cognitive test. The steeper slope in the rightmost panel suggests a stronger negative relationship between earlier pTau217 and downstream PACC-3.

**Table 1.** Demographics and background characteristics of primary analysis sample.

| Variable | Value |
| --- | --- |
| Number of participants | 165 |
| Number of plasma observations per participant, median (range) | 3 (1-5) |
| Age at first plasma, mean (SD) | 62.94 (6.06) |
| Age at last plasma, mean (SD) | 68.22 (6.10) |
| Years of plasma follow-up, mean (SD) | 5.28 (1.62) |
| Female, N (%) | 108 (65%) |
| White, N (%) | 158 (96%) |
| Black, N (%) | 4 (2%) |
| Native American, N (%) | 3 (2%) |
| Cognitively unimpaired-stable at first plasma, N (%) | 131 (79%) |
| Cognitively unimpaired-declining at first plasma, N (%) | 34 (21%) |
| Cognitively unimpaired-stable at last plasma, N (%) | 134 (81%) |
| Cognitively unimpaired-declining at last plasma, N (%) | 23 (14%) |
| MCI at last plasma, N (%) | 7 (4%) |
| Dementia at last plasma, N (%) | 1 (1%) |
| PiB- at last plasma, N (%) | 95 (58%) |
| PiB+ at last plasma, N (%) | 70 (42%) |
| MK- at last plasma, N (%) | 111 (67%) |
| MK+ MTL only at last plasma, N (%) | 6 (4%) |
| MK+ Neo only at last plasma, N (%) | 2 (1%) |
| MK+ MTL+Neo at last plasma, N (%) | 26 (16%) |
| MK+ missing, N (%) | 20 (12%) |

### Longitudinal pTau_217_ trajectories

Individual pTau_217_ trajectories are displayed by age in Figure 1A, and by GBTM-DVR in Figure 1B (secondary x-axis: centiloid conversions, at top). A strong relationship with MK-6240 PET is evident: those with tau in both medial temporal and neocortical regions appear distinct from other groups. The improved alignment in Figure 1B supports strong correspondence between plasma pTau_217_ and PiB PET.

The mixed effects models relating pTau_217_ levels to GBTM-DVR are shown in Table 2A. GBTM-DVR was a strong predictor of measured pTau_217_ (β̂_DVR_ = 0.83, *Ω*^2^ = 0.64). Intraclass correlations were moderate. Together, the results indicate good test-retest reliability and high sensitivity to true underlying change. Results of sensitivity analyses were substantially similar.

**Table 2.** Results of linear mixed effect models of pTau_217_. Each model included a per-participant random intercept. For each, sensitivity analysis 1 included observations from eight additional participants who were either cognitively unimpaired or missing a cognitive diagnosis at first available plasma draw (N=7) or whose pTau_217_ values were highly influential (N=1), and sensitivity analysis 2 excluded six single observations on five participants for which measured pTau_217_ values fell below the lower limit of detection. (A) pTau_217_ as a function of GBTM-DVR. (B) pTau_217_ as a function of age, moderated by binary PiB status.

| (2A)<br>*Predictors* | Primary set |  |  | Sensitivity set 1 |  |  | Sensitivity set 2 |  |  |
| --- | --- | --- | --- | --- | --- | --- | --- | --- | --- |
|  | *Estimates* | *CI* | *p* | *Estimates* | *CI* | *p* | *Estimates* | *CI* | *p* |
| Intercept | 0.00 | -0.08 – 0.08 | 0.968 | 0.00 | -0.12 – 0.12 | 0.972 | 0.00 | -0.08 – 0.08 | 0.984 |
| GBTM-DVR | 0.83 | 0.75 – 0.90 | **<0.001** | 0.46 | 0.35 – 0.56 | **<0.001** | 0.83 | 0.75 – 0.90 | **<0.001** |
| **Random Effects** |  |  |  |  |  |  |  |  |  |
| $\sigma^2$ | 0.12 | | | 0.32 | | | 0.12 | | |
| $\tau_{00}$ | 0.25 | Reggieid | | 0.52 | Reggieid | | 0.25 | Reggieid | |
| ICC | 0.67 |  |  | 0.62 |  |  | 0.67 |  |  |
| N | 165 | Reggieid |  | 173 | Reggieid |  | 165 | Reggieid |  |
| Observations | 515 |  |  | 530 |  |  | 509 |  |  |
| Marginal R<sup>2</sup> / Conditional R<sup>2</sup> | 0.648 / 0.885 |  |  | 0.200 / 0.695 |  |  | 0.649 / 0.885 |  |  |

| (2B)<br>*Predictors* | Primary set |  |  | Sensitivity set 1 |  |  | Sensitivity set 2 |  |  |
| --- | --- | --- | --- | --- | --- | --- | --- | --- | --- |
|  | *Estimates* | *CI* | *p* | *Estimates* | *CI* | *p* | *Estimates* | *CI* | *p* |
| Intercept | 0.23 | 0.20 – 0.27 | **<0.001** | 0.27 | 0.21 – 0.34 | **<0.001** | 0.23 | 0.20 – 0.27 | **<0.001** |
| Amyloid positivity | 0.19 | 0.13 – 0.24 | **<0.001** | 0.16 | 0.06 – 0.26 | **0.002** | 0.19 | 0.13 – 0.24 | **<0.001** |
| Age [centered], linear | 0.00 | -0.00 – 0.00 | 0.337 | 0.00 | -0.00 – 0.01 | 0.267 | 0.00 | -0.00 – 0.00 | 0.350 |
| Amyloid positivity x Age [centered], linear | 0.02 | 0.02 – 0.03 | **<0.001** | 0.01 | 0.00 – 0.03 | **0.021** | 0.02 | 0.02 – 0.03 | **<0.001** |
| **Random Effects** |  |  |  |  |  |  |  |  |  |
| $\sigma^2$ | 0.01 | | | 0.05 | | | 0.01 | | |
| $\tau_{00}$ | 0.03 | Reggieid | | 0.09 | Reggieid | | 0.03 | Reggieid | |
| ICC | 0.81 |  |  | 0.66 |  |  | 0.81 |  |  |
| N | 165 | Reggieid |  | 173 | Reggieid |  | 165 | Reggieid |  |
| Observations | 515 |  |  | 530 |  |  | 509 |  |  |
| Marginal R<sup>2</sup> / Conditional R<sup>2</sup> | 0.355 / 0.879 |  |  | 0.093 / 0.695 |  |  | 0.356 / 0.878 |  |  |

The mixed effects models relating pTau_217_ levels to age, with binary PiB status (PiB DVR > 1.19) as a moderator, are shown in Table 2B. A mid-sized, significant age by amyloid status interaction was observed such that levels of pTau_217_ increased with age only in PiB+ participants, whereas in PiB- participants, the age-related slope estimate was indistinguishable from zero (*β̂*_PiB×age_ = 0.021; *β̂*_PiB-_ = 0.0016, *β̂*_PiB+_ = 0.022; *Ω*^2^ = 0.11). One participant with high pTau_217_ levels continued to be highly influential in this model. The model fit to the primary dataset is shown in Figure 1C. Results of sensitivity analyses were similar.

### pTau_217_ threshold estimation

Boxplots and ROC curves relating pTau_217_ to binary PiB and MTL+neocortical MK- 6240 are shown in Figure 1D-G. Correspondence was high for both PiB positivity thresholds (early positivity (DVR > 1.16), AUC=0.90, PPV=0.58, NPV=0.94; late positivity (DVR > 1.19), AUC=0.91, PPV=0.58, NPV=0.94), as well as for MTL+neocortical MK-6240 positivity (AUC=0.95, PPV=1.00, NPV=0.98). Estimated thresholds were lower for amyloid than tau (early PiB positivity: 0.27; late PiB positivity: 0.27; MTL+neocortical MK-6240 positivity: 0.37). Our robust norms threshold approach identified a higher pTau_217_ positivity boundary of 0.37. Analyses on the sensitivity datasets were similar (see eFigures 1 and 2).

When adjudicating both the late PiB positivity, ROC-based threshold and the robust norms threshold against the ground truth of concurrent GBTM-DVR > 1.19, the ROC-based threshold was more sensitive, but less specific (ROC threshold: sensitivity = 0.91, specificity = 0.75; robust norms threshold: sensitivity = 0.7, specificity = 0.96). Among individuals having ≥ 1 plasma pTau_217_ observation between these thresholds (N=70), half were PiB+ at their last PET scan (N=36).

### Associations with longitudinal cognition

Figure 2A illustrates the mixed effects model relating baseline pTau_217_ levels to PACC-3 trajectories. We observed a mid-sized, significant age by pTau_217_ interaction (*β̂*_pTau217×age_ = -0.075, *Ω*^2^ = 0.10). Lower baseline pTau levels were associated with a flatter cognitive trajectory, whereas moderate and higher levels were linked to faster decline. According to this model, two women with average education, literacy, and prior exposure, but with 10th and 90th percentile levels of plasma pTau_217_, would be expected to decline to a PACC-3 score of z = -1.5 by approximately ages 91.5 and 74.2, respectively; for similar men the expected ages are 86.4 and 71.7, respectively. A likelihood ratio test comparing this model to a covariates-only version indicated better performance when including pTau_217_ (χ^2^(3) = 70.7, p < .0001). Model summaries are shown in Table 3.

**Table 3.**
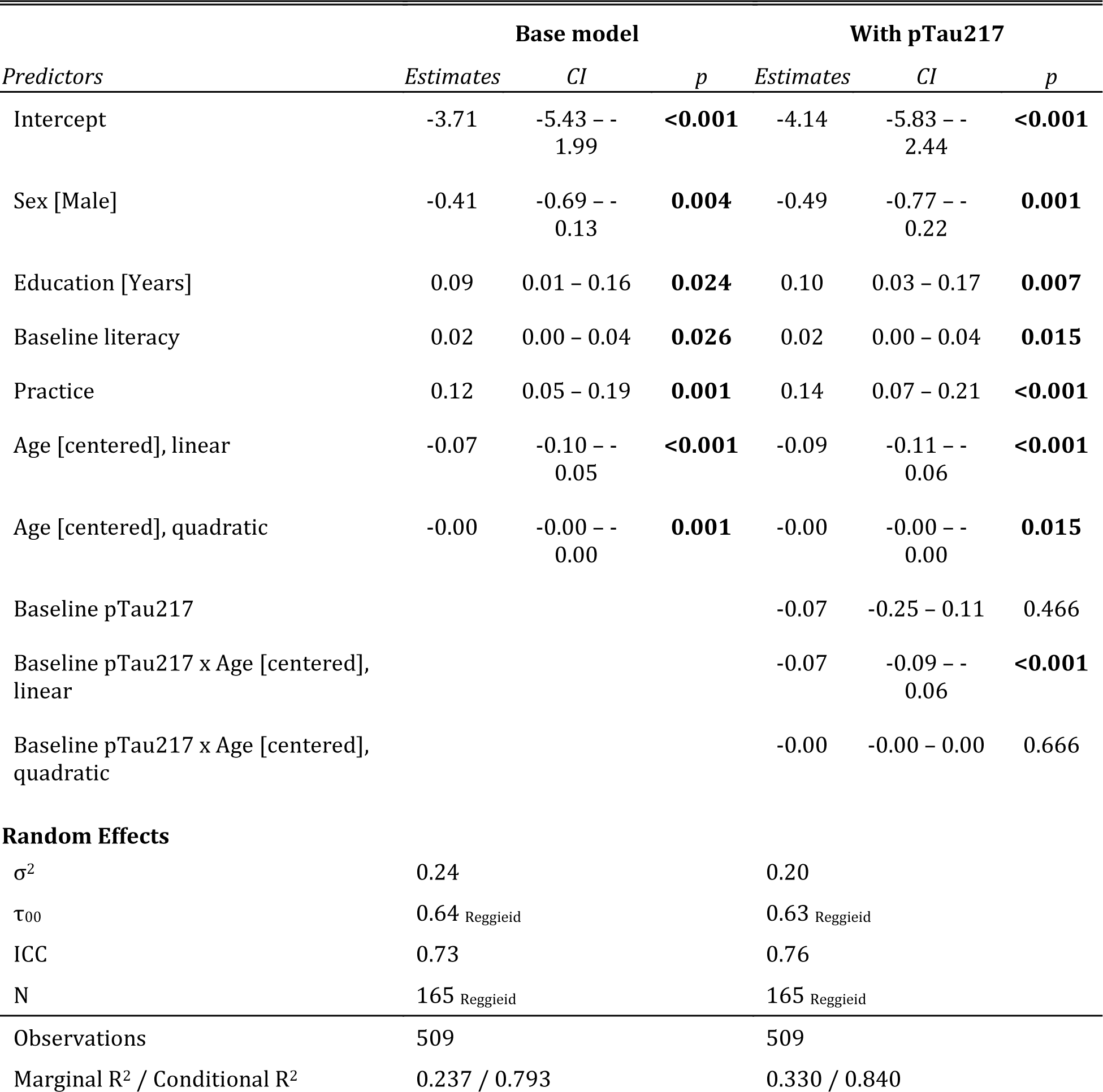
Results of linear mixed effect models of PACC-3 as a function of (1) covariates only (sex, education, baseline literacy, and age); (2) covariates plus baseline pTau_217_ and its interaction with age. In both models, age was modeled as a second-degrees polynomial. Models included a per-participant random intercept.

Sensitivity analyses were similar (see eTable 1 and eFigures 3 and 4).

Exploratory lagged repeated measures correlations on a subset of individuals with at least 4 observations (N=46) suggested weak within-person correspondence between pTau_217_ and concurrent cognition (*r*_*lag=0*_ = -0.130), but stronger within-person relationships when a lag of 1 (*r*_*lag=1*_ = -0.550) or 2 visits (*r*_*lag=2*_ = -0.670; Figure 2B-D) was imposed. This pattern held when repeated measures correlation was performed instead after age had been partialed out from both pTau_217_ and cognitive scores (*r*_*lag*_ = -0.0720; *r*_*lag=1*_ = -0.500; *r*_*lag=2*_ = -0.630). In comparison, relationships with modeled PiB DVR at the same lags were weaker (*r*_*gbtm.lag=0*_ = -0.0230; _*gbtm.lag=1*_ = -0.410; _*gbtm.lag=2*_ = -0.540).

## Discussion

We characterized the temporal dynamics of Lilly-MSD pTau_217_ in a cohort of late-middle-aged adults without baseline clinical cognitive impairment at a mean age of 63. We observed a strong relationship between brain amyloid positivity and pTau_217_, with plasma biomarker trajectories rising with age only in A+ individuals. This is similar to a recent report from BioFINDER^23^ in a sample approximately ten years older. We further observed strong relationships between pTau_217_ trajectories and brain tau as measured via MK-6240: those with extensive tau deposition, all PiB+, exhibited increasing plasma levels of pTau_217_. This resembles recent findings in AIBL of a moderately strong cross-sectional correlation between pTau_217_ + and meta-temporal and mesial temporal MK-6240 SUVR in A+ older adults.^11^ However, in that analysis, the correlation was weaker in a subset of CU participants, whereas in the present analysis, which includes only CU individuals and again features a cohort ten years younger, the relationship is strong.

Plasma biomarkers have potential for prescreening AD biomarker positive participants in clinical trials.^3^ To that end, our results are encouraging, as we observed strong relationships between plasma pTau_217_ and concurrent brain imaging biomarkers of AD, with an AUC of approximately 0.91 for identifying PiB+ participants, and 0.95 for identifying those who were MK+. These values are similar to those seen for the easier task of discriminating AD A*β*^+^ from CU A*β*^−^ groups, and are high compared to other reports describing cognitively unimpaired elderly groups in AIBL,^11^ MCSA,^24^ and BioFINDER.^46^ With our threshold for predicting PiB+, the PPV of pTau_217_ was 0.58, which would reduce the number needed to screen to obtain a full sample. However, for other purposes, a more conservative threshold might be preferable. For MK, in contrast, the PPV of 1 and NPV of 0.98 are likely overestimates, but suggest this threshold may work well for many purposes, in principle, in populations with prevalence close to our estimates.

Our two analyses relating baseline pTau_217_ to PACC-3 scores were complementary, each suggesting important longitudinal relationships between this plasma biomarker and cognition. In our primary analysis, the pTau_217_ by age interaction suggests those with higher baseline biomarker levels evince worse cognitive trajectories with age than do those with lower biomarker levels, whose cognitive trajectories appear flat. In our exploratory analysis using repeated measures correlation, although the within-person relationship between biomarker levels and concurrent cognitive performance is weak, by modeling a delayed effect using a lagged correlation, a robust negative relationship emerges. Although similar relationships have been found in older groups, our report establishes such relationships with biomarkers measured in late midlife.^3, 23, 24^ Given the interest in establishing valid surrogate outcomes for AD pharmaceutical research,^47^ our findings may inform the trial design in which the fitness of plasma pTau_217_ for that purpose is evaluated.

### Limitations

The chief limitation of the present analysis is our small, racially-homogenous sample, drawn from a cohort that is convenience- and not population-based.^27^ The complexities of bringing plasma assays into use with heterogenous clinical populations should not be discounted. However, recent work in WHICAP suggests relatively good concordance between pTau_217_ and clinical status, and no evident demographic biases.^48^

Future directions include assaying our extensive back-catalog of plasma and expanding our existing cohort with a more diverse group of research participants.

## Conclusion

In this report, we extend previous findings of strong relationships between plasma levels of pTau_217_, concurrent PET AD biomarkers, and prospective cognition to a preclinical dataset. These findings have strong implications for early detection, which is prerequisite for several major goals of AD research: understanding susceptibility and resilience factors that underlie prognosis; designing better primary and secondary AD prevention trials; and determining the relative timing and impact of AD and co-occurring pathologies on cognitive decline.

## Supporting information

STROBE checklist

## Data Availability

All data produced in the present study are available upon reasonable request to the authors.

## Acknowledgements and Disclosures

OH has acquired research support (for the institution) from ADx, AVID Radiopharmaceuticals, Biogen, Eli Lilly, Eisai, Fujirebio, GE Healthcare, Pfizer, and Roche. In the past 2 years, he has received consultancy/speaker fees from AC Immune, Amylyx, Alzpath, BioArctic, Biogen, Cerveau, Fujirebio, Genentech, Novartis, Roche, and Siemens. SCJ has served as a consultant to Eisai and Roche Diagnostics, has received an equipment grant from Roche Diagnostics, and has received support (sponsoring of an observational study and provision of precursor for tau imaging) from Cerveau Technologies. Authors EMJ, SJ, KAC, RLK, LD, NAC, NMC, KJH, BTC, and TJB have nothing to disclose.

Work at the University of Wisconsin was supported by NIH R01AG027161 (Johnson), NIH RO1AG021155 (Johnson), AARF 19614533 (Betthauser), S10 OD025245-01 (Christian) and the University of Wisconsin Institute for Clinical and Translational Research NIH UL1TR002375 (Cody). We extend our deepest thanks to the WRAP participants and staff for their invaluable contributions to the study.

Work at Lund University was supported by the Swedish Research Council (2016- 00906), the Knut and Alice Wallenberg foundation (2017-0383), the Marianne and Marcus Wallenberg foundation (2015.0125), the Strategic Research Area MultiPark (Multidisciplinary Research in Parkinson’s disease) at Lund University, the Swedish Alzheimer Foundation (AF-939932), the Swedish Brain Foundation (FO2021-0293), The Parkinson foundation of Sweden (1280/20), the Konung Gustaf V:s och Drottning Victorias Frimurarestiftelse, the Skåne University Hospital Foundation (2020-O000028), Regionalt Forskningsstöd (2020-0314) and the Swedish federal government under the ALF agreement (2018-Projekt0279).

## eFigure Captions

**eFigure 1.**
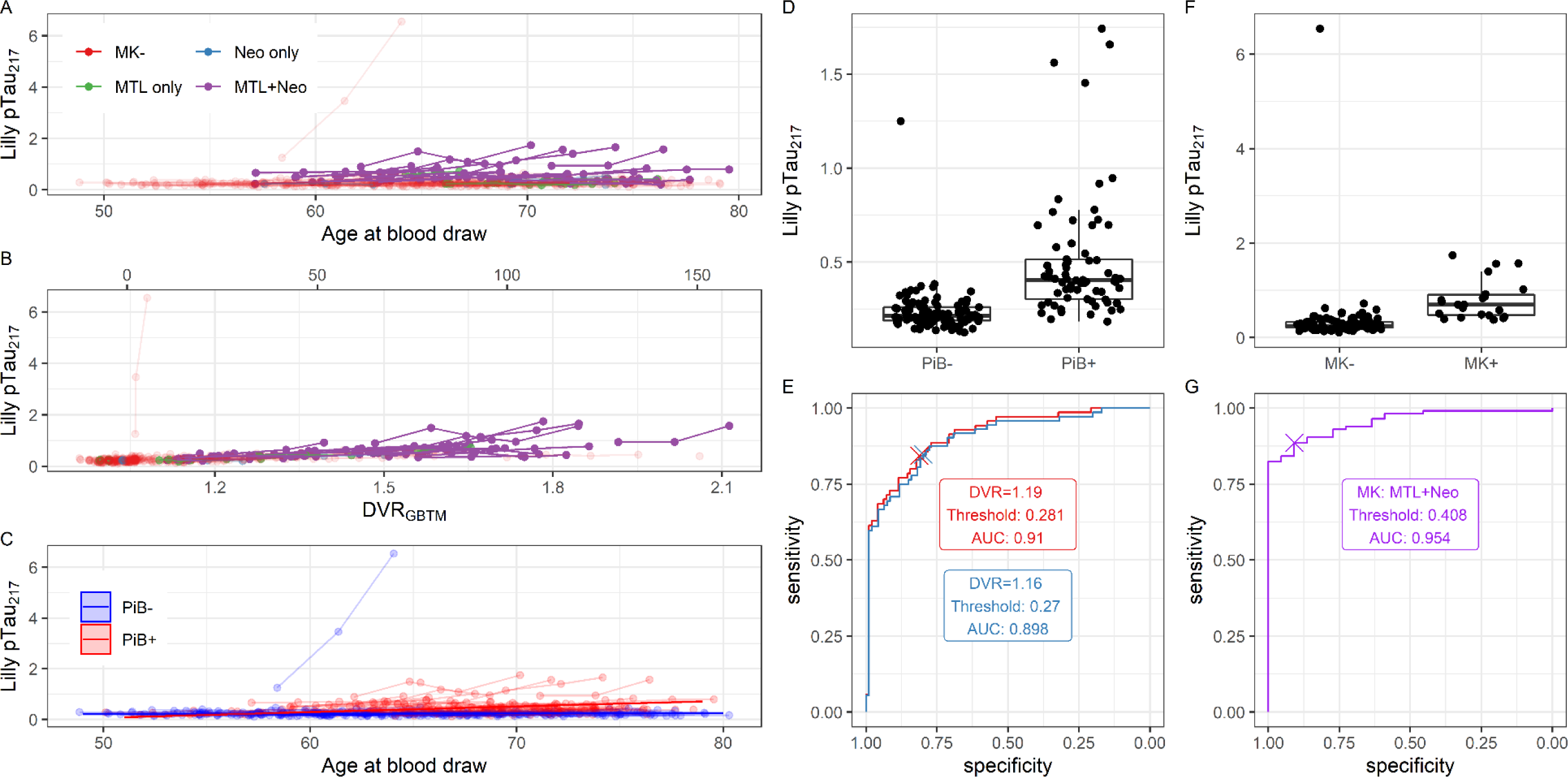
Longitudinal plasma pTau_217_, sensitivity set 1 (including 6 participants who were cognitively impaired at baseline; one with no baseline diagnosis; and one influential pTau_217_ observation). Observations from a single participant are shown with connected edges. (A) Plasma pTau_217_ as a function of age at blood draw. Color indicates the extent of tau burden as indicated on tau PET. (B) Plasma pTau_217_ as a function of estimated PiB DVR at the time of plasma acquisition. Color indicates the extent of tau burden as indicated on tau PET. (C) Plasma pTau_217_ as a function of age at blood draw. Color indicates amyloid PET positivity. Lines with shaded confidence bands represent slope estimates from a linear mixed effects model of pTau_217_ as a function of the interaction of age and amyloid positivity. (D) Distribution of pTau_217_ among PiB- and PiB+ participants. (E) Receiver-operator characteristic (ROC) curve relating pTau_217_ to binary PiB status. Two positivity thresholds were considered for PiB: global DVR > 1.19 (red) and global DVR >1.16 (blue). (F) Distribution of pTau_217_ among MK- and MK+ participants. (G) ROC curve relating pTau_217_ to binary MK status. Scans were marked as MK+ if tracer binding was evident in both medial temporal lobe and neocortex, and MK- otherwise.

**eFigure 2.**
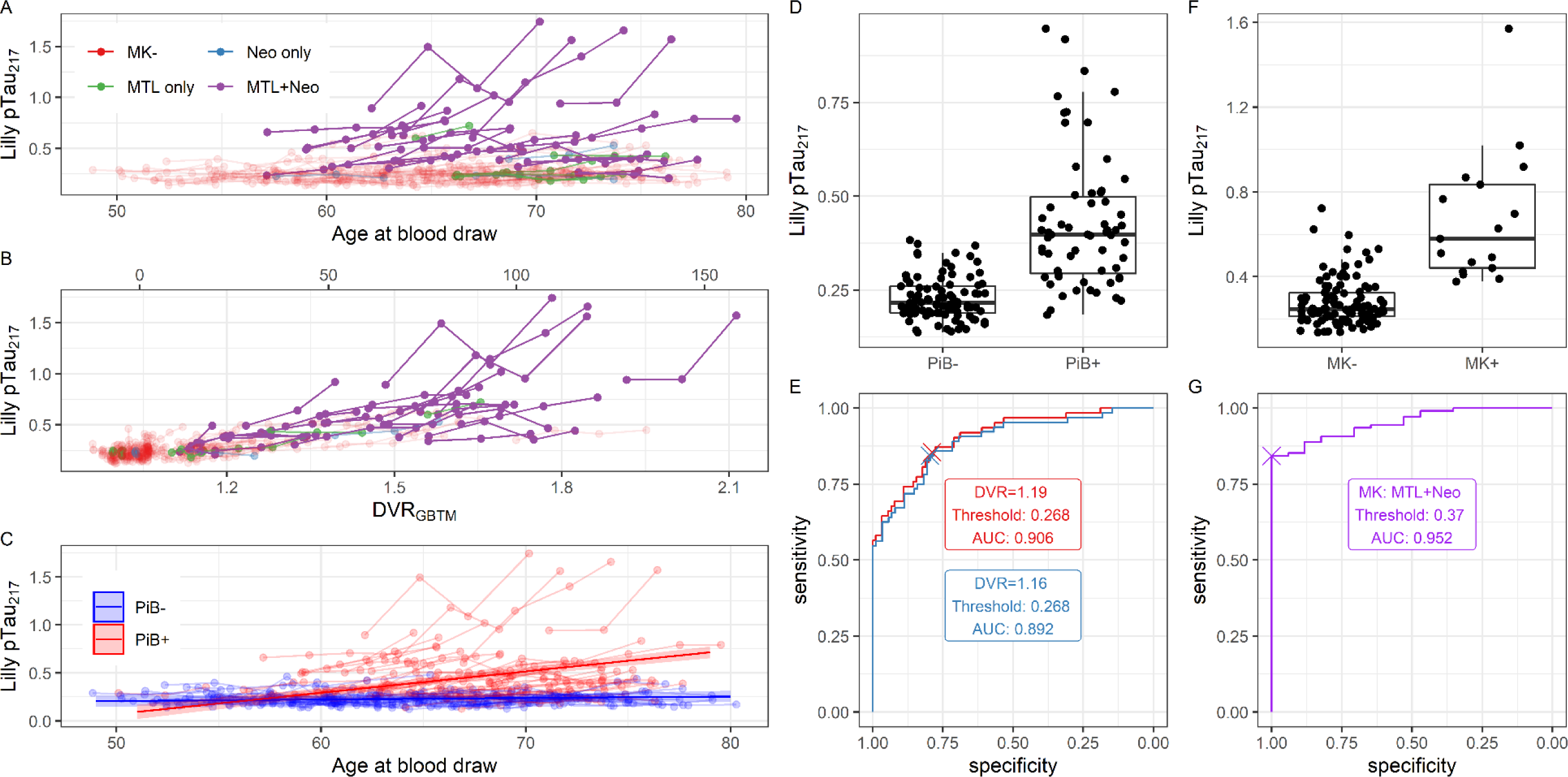
Longitudinal plasma pTau_217_, sensitivity set 2 (excluding 6 observations from 5 participants in which the pTau_217_ value fell below the lower limit of detection). Observations from a single participant are shown with connected edges. (A) Plasma pTau_217_ as a function of age at blood draw. Color indicates the extent of tau burden as indicated on tau PET. (B) Plasma pTau_217_ as a function of estimated PiB DVR at the time of plasma acquisition. Color indicates the extent of tau burden as indicated on tau PET. (C) Plasma pTau_217_ as a function of age at blood draw. Color indicates amyloid PET positivity. Lines with shaded confidence bands represent slope estimates from a linear mixed effects model of pTau_217_ as a function of the interaction of age and amyloid positivity. (D) Distribution of pTau_217_ among PiB- and PiB+ participants. (E) Receiver-operator characteristic (ROC) curve relating pTau_217_ to binary PiB status. Two positivity thresholds were considered for PiB: global DVR > 1.19 (red) and global DVR > 1.16 (blue). (F) Distribution of pTau_217_ among MK- and MK+ participants. (G) ROC curve relating pTau_217_ to binary MK status. Scans were marked as MK+ if tracer binding was evident in both medial temporal lobe and neocortex, and MK- otherwise.

**eFigure 3.**
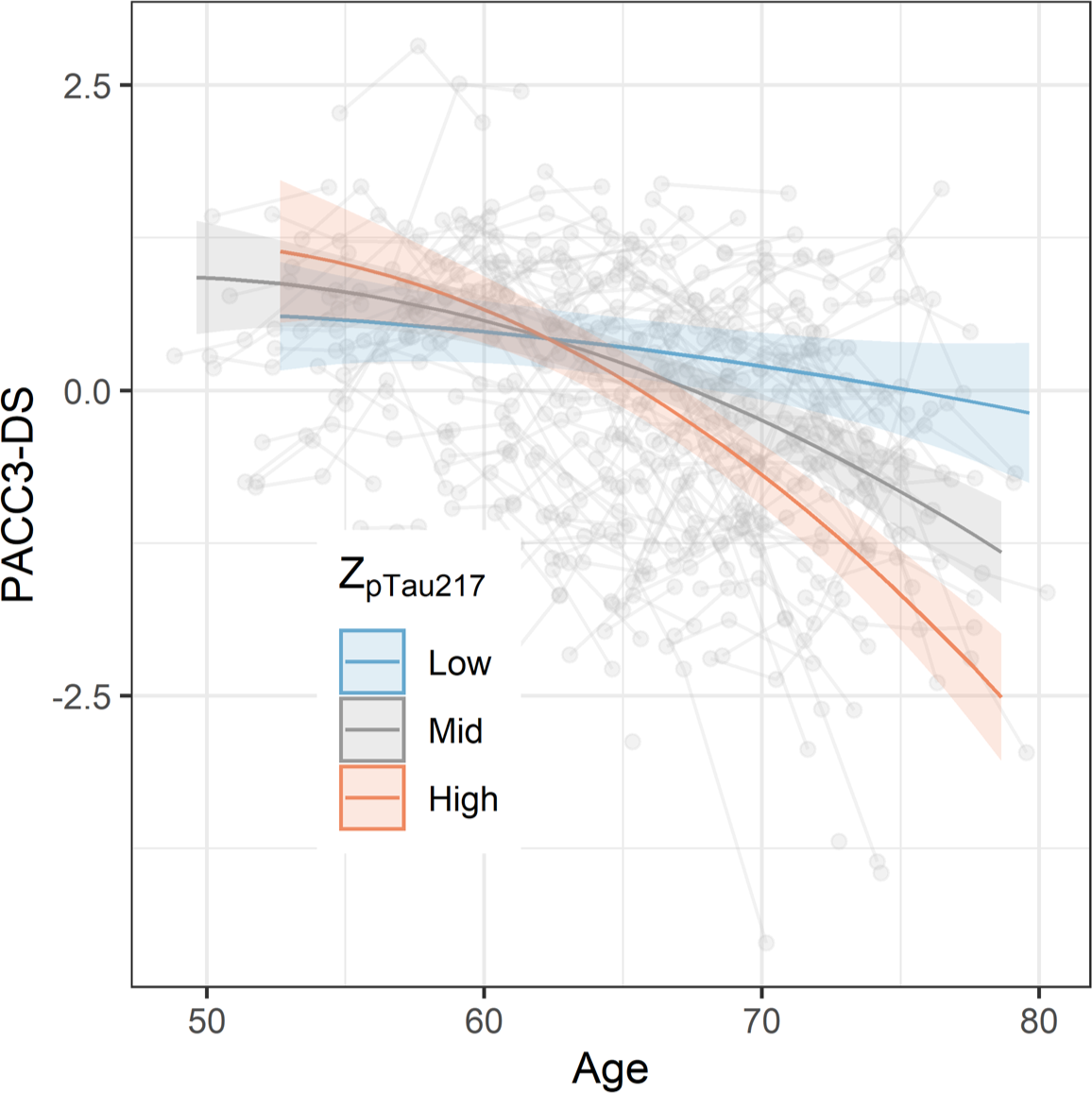
Relationships between pTau_217_ and longitudinal cognition, sensitivity set 1 (including 6 participants who were cognitively impaired at baseline; one with no baseline diagnosis; and one influential pTau_217_ observation). Global cognition (PACC-3) as a function of age and baseline pTau_217_ level. Low, medium, and high pTau_217_ values reflect the 10th, 50th, and 90th sample percentiles.

**eFigure 4.**
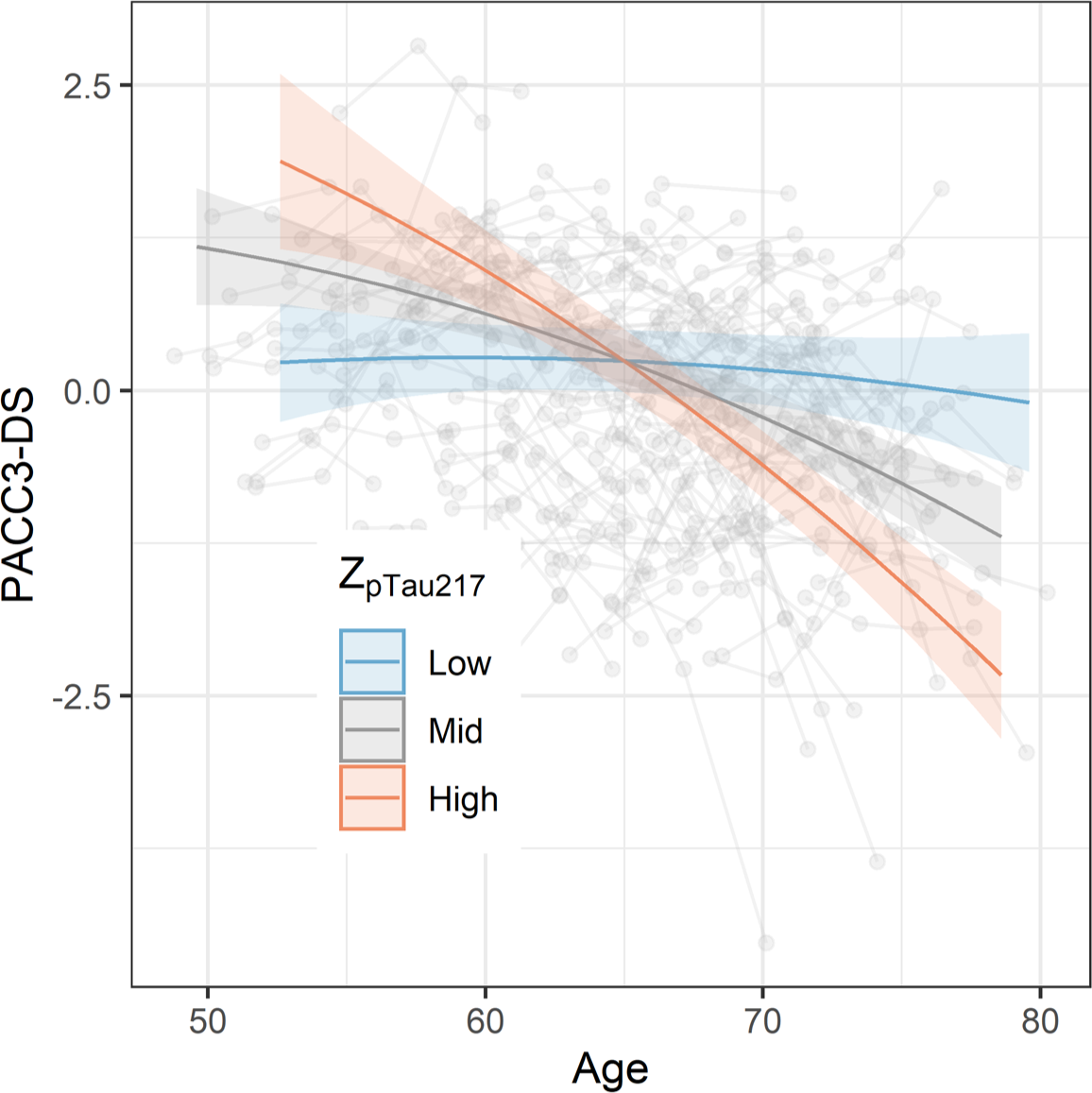
Relationships between pTau_217_ and longitudinal cognition, sensitivity set 2 (excluding 6 observations from 5 participants in which the pTau_217_ value fell below the lower limit of detection). Global cognition (PACC-3) as a function of age and baseline pTau_217_ level. Low, medium, and high pTau_217_ values reflect the 10th, 50th, and 90th sample percentiles.

**Supplement eTable 1.**
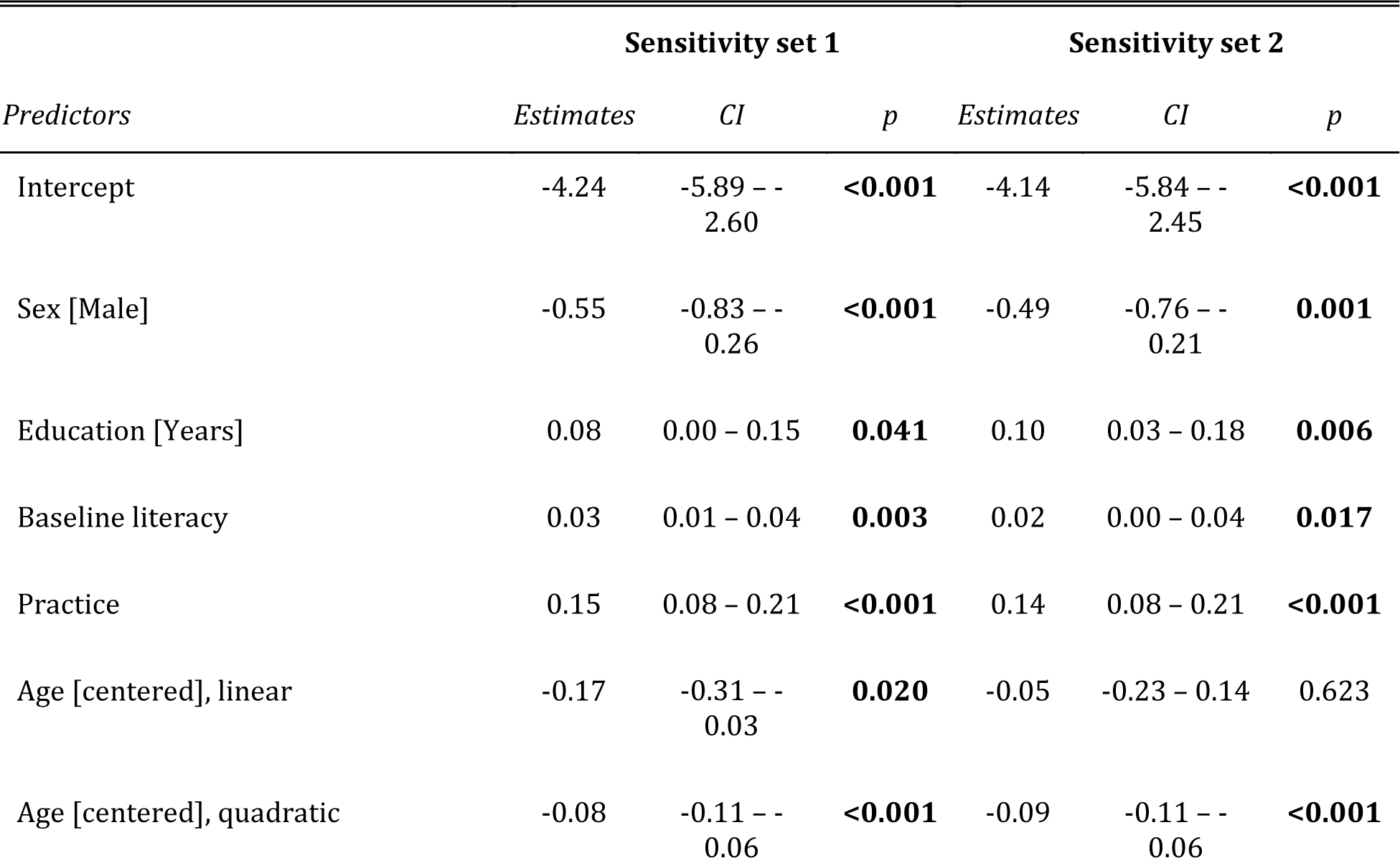
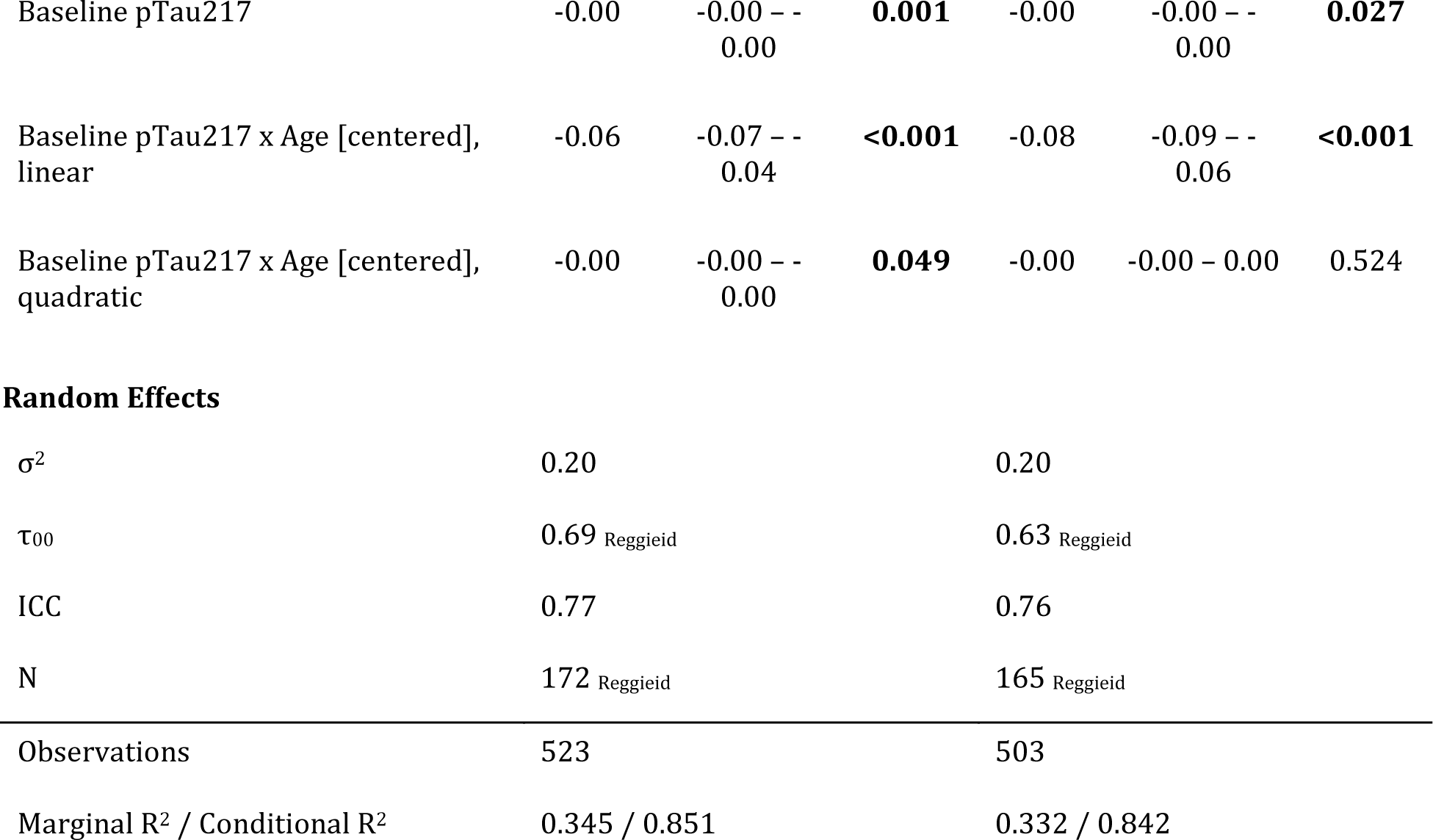
Results of linear mixed effect models of PACC-3 as a function of covariates plus baseline plasma pTau_217_ and its interaction with age, in two sensitivity sets. The first sensitivity set includes the primary analysis set plus eight additional participants who were cognitively impaired at first plasma draw (N=6), who did not have a consensus diagnosis at this visit (N=1) or who had extreme values of pTau_217_ (N=1). The second sensitivity set includes the primary analysis set, minus 6 observations on 5 participants for which the observed pTau_217_ values were below the lower limit of detection. In both models, age was modeled as a second-degrees polynomial. Models included a per-participant random intercept.

| *Predictors* | **Sensitivity set 1** |  |  | **Sensitivity set 2** |  |  |
| --- | --- | --- | --- | --- | --- | --- |
|  | *Estimates* | *CI* | *p* | *Estimates* | *CI* | *p* |
| Intercept | -4.24 | -5.89 – -2.60 | **<0.001** | -4.14 | -5.84 – -2.45 | **<0.001** |
| Sex [Male] | -0.55 | -0.83 – -0.26 | **<0.001** | -0.49 | -0.76 – -0.21 | **0.001** |
| Education [Years] | 0.08 | 0.00 – 0.15 | **0.041** | 0.10 | 0.03 – 0.18 | **0.006** |
| Baseline literacy | 0.03 | 0.01 – 0.04 | **0.003** | 0.02 | 0.00 – 0.04 | **0.017** |
| Practice | 0.15 | 0.08 – 0.21 | **<0.001** | 0.14 | 0.08 – 0.21 | **<0.001** |
| Age [centered], linear | -0.17 | -0.31 – -0.03 | **0.020** | -0.05 | -0.23 – 0.14 | 0.623 |
| Age [centered], quadratic | -0.08 | -0.11 – -0.06 | **<0.001** | -0.09 | -0.11 – -0.06 | **<0.001** |
| Baseline pTau217 | -0.00 | -0.00 – -<br>0.00 | **0.001** | -0.00 | -0.00 – -<br>0.00 | **0.027** |
| Baseline pTau217 x Age [centered],<br>linear | -0.06 | -0.07 – -<br>0.04 | **<0.001** | -0.08 | -0.09 – -<br>0.06 | **<0.001** |
| Baseline pTau217 x Age [centered],<br>quadratic | -0.00 | -0.00 – -<br>0.00 | **0.049** | -0.00 | -0.00 – 0.00 | 0.524 |
| **Random Effects** |  |  |  |  |  |  |
| $\sigma^2$ | 0.20 | | | 0.20 | | |
| $\tau_{00}$ | 0.69 | Reggieid | | 0.63 | Reggieid | |
| ICC | 0.77 |  |  | 0.76 |  |  |
| N | 172 | Reggieid |  | 165 | Reggieid |  |
| Observations | 523 |  |  | 503 |  |  |
| Marginal R<sup>2</sup> / Conditional R<sup>2</sup> | 0.345 / 0.851 |  |  | 0.332 / 0.842 |  |  |

## Notes

### Author Declarations

The Health Sciences IRB of University of Wisconsin-Madison gave ethical approval for this work

## References

1. Hansson O. Biomarkers for neurodegenerative diseases. Nature Medicine. 2021;27(6):954–963. doi:10.1038/s41591-021-01382-x

2. Leuzy A, Mattsson-Carlgren N, Palmqvist S, Janelidze S, Dage JL, Hansson O. Blood-based biomarkers for Alzheimer’s disease. EMBO molecular medicine. 2022;14(1):e14408. doi:10.15252/emmm.202114408

3. Cullen NC, Leuzy A, Janelidze S, et al. Plasma biomarkers of Alzheimer’s disease predict cognitive decline and could improve clinical trials in the cognitively unimpaired elderly. January 2021:2021.01.22.21250293. doi:10.1101/2021.01.22.21250293

4. Bateman RJ, Blennow K, Doody R, et al. Plasma Biomarkers of AD Emerging as Essential Tools for Drug Development: An EU/US CTAD Task Force Report. The Journal of Prevention of Alzheimer’s Disease. 2019;6(3):169–173. doi:10.14283/jpad.2019.21

5. West T, Kirmess KM, Meyer MR, et al. A blood-based diagnostic test incorporating plasma A*β*42/40 ratio, ApoE proteotype, and age accurately identifies brain amyloid status: Findings from a multi cohort validity analysis. Molecular Neurodegeneration. 2021;16(1):30. doi:10.1186/s13024-021-00451-6

6. Ovod V, Ramsey KN, Mawuenyega KG, et al. Amyloid *β* concentrations and stable isotope labeling kinetics of human plasma specific to central nervous system amyloidosis. Alzheimer’s & Dementia: The Journal of the Alzheimer’s Association. 2017;13(8):841–849. doi:10.1016/j.jalz.2017.06.2266

7. Nakamura A, Kaneko N, Villemagne VL, et al. High performance plasma amyloid-*β* biomarkers for Alzheimer’s disease. Nature. 2018;554(7691):249-254. doi:10.1038/nature25456

8. Palmqvist S, Janelidze S, Stomrud E, et al. Performance of Fully Automated Plasma Assays as Screening Tests for Alzheimer Disease-Related *β*-Amyloid Status. JAMA neurology. 2019;76(9):1060–1069. doi:10.1001/jamaneurol.2019.1632

9. Palmqvist S, Janelidze S, Quiroz YT, et al. Discriminative Accuracy of Plasma Phospho-tau217 for Alzheimer Disease vs Other Neurodegenerative Disorders. JAMA. 2020;324(8):1–11. doi:10.1001/jama.2020.12134

10. Karikari TK, Pascoal TA, Ashton NJ, et al. Blood phosphorylated tau 181 as a biomarker for Alzheimer’s disease: A diagnostic performance and prediction modelling study using data from four prospective cohorts. The Lancet Neurology. 2020;19(5):422–433. doi:10.1016/S1474-4422(20)30071-5

11. Doré V, Doecke JD, Saad ZS, et al. Plasma P217+tau versus NAV4694 amyloid and MK6240 tau PET across the Alzheimer’s continuum. *Alzheimer’s & Dementia (Amsterdam*, Netherlands). 2022;14(1):e12307. doi:10.1002/dad2.12307

12. Mattsson N, Cullen NC, Andreasson U, Zetterberg H, Blennow K. Association Between Longitudinal Plasma Neurofilament Light and Neurodegeneration in Patients With Alzheimer Disease. JAMA neurology. 2019;76(7):791–799. doi:10.1001/jamaneurol.2019.0765

13. Ashton NJ, Janelidze S, Al Khleifat A, et al. A multicentre validation study of the diagnostic value of plasma neurofilament light. Nature Communications. 2021;12(1):3400. doi:10.1038/s41467-021-23620-z

14. Elahi FM, Casaletto KB, La Joie R, et al. Plasma biomarkers of astrocytic and neuronal dysfunction in early- and late-onset Alzheimer’s disease. Alzheimer’s & Dementia: The Journal of the Alzheimer’s Association. 2020;16(4):681–695. doi:10.1016/j.jalz.2019.09.004

15. Pereira JB, Janelidze S, Smith R, et al. Plasma GFAP is an early marker of amyloid-*β* but not tau pathology in Alzheimer’s disease. Brain: A Journal of Neurology. 2021;144(11):3505–3516. doi:10.1093/brain/awab223

16. Benedet AL, Milà-Alomà M, Vrillon A, et al. Differences Between Plasma and Cerebrospinal Fluid Glial Fibrillary Acidic Protein Levels Across the Alzheimer Disease Continuum. JAMA neurology. 2021;78(12):1471–1483. doi:10.1001/jamaneurol.2021.3671

17. Barthélemy NR, Li Y, Joseph-Mathurin N, et al. A soluble phosphorylated tau signature links tau, amyloid and the evolution of stages of dominantly inherited Alzheimer’s disease. Nature Medicine. 2020;26(3):398–407. doi:10.1038/s41591-020-0781-z

18. Karikari TK, Emeršič A, Vrillon A, et al. Head-to-head comparison of clinical performance of CSF phospho-tau T181 and T217 biomarkers for Alzheimer’s disease diagnosis. Alzheimer’s & Dementia. 2021;17(5):755–767. doi:10.1002/alz.12236

19. Thijssen EH, La Joie R, Strom A, et al. Plasma phosphorylated tau 217 and phosphorylated tau 181 as biomarkers in Alzheimer’s disease and frontotemporal lobar degeneration: A retrospective diagnostic performance study. The Lancet Neurology. 2021;20(9):739–752. doi:10.1016/S1474-4422(21)00214-3

20. Wennström M, Janelidze S, Nilsson KPR, et al. Cellular localization of p-Tau217 in brain and its association with p-Tau217 plasma levels. Acta Neuropathologica Communications. 2022;10(1):3. doi:10.1186/s40478-021-01307-2

21. Mattsson-Carlgren N, Janelidze S, Bateman RJ, et al. Soluble P-tau217 reflects amyloid and tau pathology and mediates the association of amyloid with tau. EMBO molecular medicine. 2021;13(6):e14022. doi:10.15252/emmm.202114022

22. Janelidze S, Berron D, Smith R, et al. Associations of Plasma Phospho-Tau217 Levels With Tau Positron Emission Tomography in Early Alzheimer Disease. JAMA neurology. 2021;78(2):149–156. doi:10.1001/jamaneurol.2020.4201

23. Mattsson-Carlgren N, Janelidze S, Palmqvist S, et al. Longitudinal plasma p-Tau217 is increased in early stages of Alzheimer’s disease. Brain: A Journal of Neurology. 2020;143(11):3234–3241. doi:10.1093/brain/awaa286

24. Mielke MM, Frank RD, Dage JL, et al. Comparison of Plasma Phosphorylated Tau Species With Amyloid and Tau Positron Emission Tomography, Neurodegeneration, Vascular Pathology, and Cognitive Outcomes. JAMA neurology. 2021;78(9):1108–1117. doi:10.1001/jamaneurol.2021.2293

25. Triana-Baltzer G, Moughadam S, Slemmon R, et al. Development and validation of a high-sensitivity assay for measuring P217+tau in plasma. *Alzheimer’s & Dementia (Amsterdam*, Netherlands). 2021;13(1):e12204. doi:10.1002/dad2.12204

26. Barthélemy NR, Horie K, Sato C, Bateman RJ. Blood plasma phosphorylated-tau isoforms track CNS change in Alzheimer’s disease. The Journal of Experimental Medicine. 2020;217(11):e20200861. doi:10.1084/jem.20200861

27. Johnson SC, Koscik RL, Jonaitis EM, et al. The Wisconsin Registry for Alzheimer’s Prevention: A review of findings and current directions. *Alzheimer’s & Dementia (Amsterdam*, Netherlands). 2018;10:130–142. doi:10.1016/j.dadm.2017.11.007

28. Janelidze S, Palmqvist S, Leuzy A, et al. Detecting amyloid positivity in early Alzheimer’s disease using combinations of plasma A*β*42/A*β*40 and p-tau. Alzheimer’s & Dementia: The Journal of the Alzheimer’s Association. 2022;18(2):283–293. doi:10.1002/alz.12395

29. Johnson SC, Christian BT, Okonkwo OC, et al. Amyloid burden and neural function in people at risk for Alzheimer’s Disease. Neurobiology of aging. 2014;35(3):576–584. doi:10.1016/j.neurobiolaging.2013.09.028

30. Betthauser TJ, Cody KA, Zammit MD, et al. In Vivo Characterization and Quantification of Neurofibrillary Tau PET Radioligand 18F-MK-6240 in Humans from Alzheimer Disease Dementia to Young Controls. *Journal of Nuclear Medicine: Official Publication*, Society of Nuclear Medicine. 2019;60(1):93–99. doi:10.2967/jnumed.118.209650

31. Racine AM, Clark LR, Berman SE, et al. Associations between Performance on an Abbreviated CogState Battery, Other Measures of Cognitive Function, and Biomarkers in People at Risk for Alzheimer’s Disease. Journal of Alzheimer’s disease : JAD. 2016;54(4):1395–1408. doi:10.3233/JAD-160528

32. Farrell ME, Jiang S, Schultz AP, et al. Defining the Lowest Threshold for Amyloid-PET to Predict Future Cognitive Decline and Amyloid Accumulation. Neurology. 2021;96(4):e619–e631. doi:10.1212/WNL.0000000000011214

33. Koscik RL, Betthauser TJ, Jonaitis EM, et al. Amyloid duration is associated with preclinical cognitive decline and tau PET. *Alzheimer’s & Dementia: Diagnosis*, Assessment & Disease Monitoring. 2020;12(1). doi:10.1002/dad2.12007

34. Betthauser TJ, Bilgel M, Koscik RL, et al. Multi-method investigation of factors influencing amyloid onset and impairment in three cohorts. December 2021:2021.12.02.21266523. doi:10.1101/2021.12.02.21266523

35. Koscik RL, La Rue A, Jonaitis EM, et al. Emergence of mild cognitive impairment in late middle-aged adults in the Wisconsin Registry for Alzheimer’s Prevention. Dementia and Geriatric Cognitive Disorders. 2014;38(1-2):16–30. doi:10.1159/000355682

36. Langhough Koscik R, Hermann BP, Allison S, et al. Validity Evidence for the Research Category, “Cognitively Unimpaired - Declining,” as a Risk Marker for Mild Cognitive Impairment and Alzheimer’s Disease. Frontiers in Aging Neuroscience. 2021;13:688478. doi:10.3389/fnagi.2021.688478

37. Jonaitis EM, Koscik RL, Clark LR, et al. Measuring longitudinal cognition: Individual tests versus composites. Alzheimer’s & Dementia: Diagnosis, Assessment & Disease Monitoring. 2019;11:74–84. doi:10.1016/j.dadm.2018.11.006

38. R Core Team. R: A Language and Environment for Statistical Computing. 2017.

39. Bates D, Mächler M, Bolker B, Walker S. Fitting linear mixed-effects models using lme4. Journal of Statistical Software. 2015;67(1):1–48. doi:10.18637/jss.v067.i01

40. Xu R. Measuring explained variation in linear mixed effects models. Statistics in Medicine. 2003;22(22):3527–3541. doi:10.1002/sim.1572

41. Ben-Shachar MS, Lüdecke D, Makowski D. effectsize: Estimation of effect size indices and standardized parameters. Journal of Open Source Software. 2020;5(56):2815. doi:10.21105/joss.02815

42. Robin X, Turck N, Hainard A, et al. pROC: An open-source package for R and S+ to analyze and compare ROC curves. BMC Bioinformatics. 2011;12:77.

43. Youden WJ. Index for rating diagnostic tests. Cancer. 1950;3(1):32–35. doi:10.1002/1097-0142(1950)3:1#x003C;32::aid-cncr2820030106>3.0.co;2-3

44. Bakdash JZ, Marusich LR. Repeated Measures Correlation. Frontiers in Psychology. 2017;8:456. doi:10.3389/fpsyg.2017.00456

45. Bakdash JZ, Marusich LR. Rmcorr: Repeated Measures Correlation.; 2021.

46. Palmqvist S, Janelidze S, Quiroz YT, et al. Discriminative Accuracy of Plasma Phospho-tau217 for Alzheimer Disease vs Other Neurodegenerative Disorders. JAMA. 2020;324(8):772–781. doi:10.1001/jama.2020.12134

47. Planche V, Villain N. US Food and Drug Administration Approval of Aducanumab-Is Amyloid Load a Valid Surrogate End Point for Alzheimer Disease Clinical Trials? JAMA neurology. 2021;78(11):1307–1308. doi:10.1001/jamaneurol.2021.3126

48. Brickman AM, Manly JJ, Honig LS, et al. Correlation of plasma and neuroimaging biomarkers in Alzheimer’s disease. Annals of Clinical and Translational Neurology. March 2022. doi:10.1002/acn3.51529

